# The humoral response to BK polyomavirus in kidney transplant recipients is dominated by IgM antibodies that use a distinct repertoire compared to IgG against the same antigen

**DOI:** 10.1101/2021.02.04.21250913

**Authors:** Nguyen Ngoc-Khanh, Gautreau-Rolland Laetitia, Devilder Marie-Claire, Fourgeux Cynthia, Sinha Debajyoti, Poschmann Jeremie, Hourmant Maryvonne, Bressollette-Bodin Céline, Saulquin Xavier, McIlroy Dorian

**Affiliations:** CRTI, UMR 1064 INSERM, Université de Nantes; CRCINA, UMR 1232 INSERM, Université de Nantes; UFR Sciences et Techniques, Université de Nantes; Service du Virologie, CHU Nantes; Service de Néphrologie-Immunologie clinique, CHU Nantes; UFR Médecine, Université de Nantes

## Abstract

The BK polyomavirus (BKPyV) persists asymptomatically in the kidney and active replication is only seen in immunosuppressed individuals, such as kidney transplant (KTx) recipients, in whom BKPyV reactivation can cause significant morbidity. KTx recipients with BKPyV reactivation mount a robust humoral response, but this often fails to clear the virus. In order to characterize the BKPyV-specific B-cell receptor (BCR) repertoire in KTx recipients, we used fluorescence-labeled BKPyV virus-like particles (VLPs) to sort with BKPyV-specific B-cells, then single-cell RNAseq to obtain paired heavy and light chain antibody sequences, and gene transcriptome data. The BCR repertoire was highly diverse in terms of both V-gene usage and clonotype diversity, with approximately 3% repertoire overlap between patients. The BKPyV-specific response was characterized by the presence of both memory IgG and memory IgM B-cells with extensive somatic hypermutation, which expressed distinct BCR repertoires within the same patient. The gene expression profile of IgG and IgM memory B-cells was highly similar, with only 19 genes, including *CD83, CD79A* and *PARP1* showing significant differential expression. These results confirm that the IgM memory B-cells are a significant component of the BKPyV-specific humoral response, and show for the first time that IgG and IgM repertoires directed against the same antigen can have significant differences.

## 2. Introduction

The BK polyomavirus PyV (BKPyV) is a typical opportunistic pathogen. Following asymptomatic primary infection during childhood, it establishes a latent infection in the kidney which appears to persist throughout life. Approximately 7% of healthy adults excrete BKPyV in the urine (Egli et al., 2009), and this proportion increases during acquired (Sundsfjord et al., 1994), or iatrogenic (Leung et al., 2001) immunosuppression. Its pathogenic potential is manifested in patients treated with allogeneic hematopoietic stem-cell transplantation (HSCT), in whom BKPyV replication can cause hemorrhagic cystitis (BKPyV-HC), and kidney transplant (KTx) recipients, in whom uncontrolled BKPyV replication can result in polyomavirus nephropathy (PyVAN) and graft loss or dysfunction. PyVAN can only be diagnosed definitively by histology, but it is correlated with DNaemia greater than 10^4^ genome copies/mL (V. Nickeleit et al., 2000), and high-level DNaemia is generally classified as presumptive PyVAN (Hirsch et al., 2019). There is currently no approved antiviral therapy with clinical efficacy against BKPyV, so presumptive or biopsy-confirmed PyVAN is managed by modulation of immunosuppressive therapy, which allows host immune responses to clear the virus (Babel et al., 2011). The virological response rate to this intervention appears to vary between centres, with recent publications reporting clearance of DNAemia in response to modulation of immunosuppression in proportions varying from 30% (Bruminhent et al., 2019; Garofalo et al., 2019) up to more than 75% (Mühlbacher et al., 2020) of PyVAN patients. In the single-centre study with the longest follow-up and the largest cohort, at least 25% of PyVAN patients had DNAemia that persisted for more than one year, despite modulation of immunosuppressive therapy (Bischof et al., 2019). Similarly, the Banff working group on PyVAN, analyzing data from nine transplant centres in Europe and North America, found that PyVAN persisted for more than 24 months in 39 of 149 (26%) patients (Volker Nickeleit et al., 2018). Importantly, recent analysis indicates that persistent PyVAN is associated with an increased risk of graft failure, and that graft loss occurs almost exclusively in patients with persistent PyVAN (Nickeleit et al., 2020).

Several previous studies have shown that the antiviral CTL response plays a key role in BKPyV clearance (Binggeli et al., 2007; Leboeuf et al., 2017; Schachtner et al., 2011), leading to the development of cellular immunotherapy for active BKPyV replication in the context of HSCT (Papadopoulou et al., 2014; Tzannou et al., 2017) and solid organ transplant (Nelson et al., 2020). With respect to the BKPyV-specific humoral response, it has been observed that the incidence of PyVAN is higher in KTx recipients with low ELISA (Bohl et al., 2008) and neutralizing antibody titres (Solis et al., 2018), and some studies and case reports have indicated that infusion of intravenous immunoglobulin (IVIG) containing BKPyV-specific neutralizing antibodies (Velay et al., 2019), can prevent active BKPyV replication in KTx recipients (Benotmane et al., 2021), and successfully treat PyVAN (Hwang et al., 2018; Matsumura et al., 2020; Piburn & Al-Akash, 2020).

However, several studies have shown that the robust humoral response following BKPyV reactivation does not always coincide with control of virus replication (Bohl et al., 2008; Chen et al., 2006; Schachtner et al., 2011). Persistent BKPyV DNaemia in the face of a strong humoral response appears to be related to the emergence of neutralization escape mutations in the virus capsid (McIlroy et al., 2020; Peretti et al., 2018), which raises the question of whether differences exist in the quality of the BKPyV-specific humoral response that might predispose some KTx recipients to neutralization escape. In particular, KTx recipients who fail to control BKPyV replication despite a strong antibody response could have a narrower BKPyV-specific B-cell receptor (BCR) repertoire, focused mainly on variable capsid regions such as the BC-loop, rather than the conserved epitopes that are recognized by broadly neutralizing antibodies described in healthy blood donors (Lindner 2019).

In the present work we aimed to characterize features of the BKPyV-specific BCR repertoire in KTx recipients who had developed a strong humoral response to the virus. By combining sorting of specific B-cells with fluorescence-labeled BKPyV virus-like particles and single cell RNAseq on the 10x Genomics platform, we were able to analyze both the BCR repertoire and gene expression profile in KTx recipients’ circulating BKPyV-specific B-cells.

## 3. Materials and Methods

### Patients and Clinical Samples

Patients included in this study were transplanted in 2011 and 2012, and had previously been included in a prospective observational study, approved by the local ethics committee and declared to the French Commission Nationale de l’Informatique et des Libertés (CNIL, n°1600141). All patients gave informed consent authorizing the use of archived urine and blood samples for research purposes. Anonymized clinical and biological data for these patients were extracted from the hospital databases. The six patients in this study were diagnosed with BKPyV reactivation based on the detection of viruria >10^7^ copies/mL correlated with an increase in serum neutralizing titre >1 log10. A total of 17 PBMC samples (two to four per patient) were collected at different timepoints (1-12 months) after peak viral load and were cryopreserved in liquid nitrogen.

### Cell culture

HEK 293TT cells, purchased from the National Cancer Institute’s Developmental Therapeutics Program (Frederick, Maryland, USA), were grown in complete DMEM High Glucose (ThermoFisher) containing 10% FBS (Dutscher), 100 U/mL penicillin, 100 μg/mL streptomycin (Dutscher), 1x Glutamax-I (ThermoFisher) and 250 µg/mL Hygromycin B (Sigma). RS cells (Evercyte, Vienna, Austria), an immortalized human renal tubule epithelial cell line, were maintained in serum-free medium OptiPRO (ThermoFisher) supplemented with 100 U/mL penicillin, 100 µg/mL streptomycin (ThermoFisher) and 1x Glutamax-I (ThermoFisher) in tissue-culture plasticware coated with 50 µg/mL Collagen I (ThermoFisher). Cells were maintained at 37°C in a humidified 5% CO_2_ incubator, and passaged at confluence by trypsinization for 10 minutes with 1x TrypLE Express (ThermoFisher).

### Plasmids

The BKPyV VP2 and VP3 expression plasmids ph2b (#32109) and ph3b (#32110), as well as the murine polyomavirus (MPyV) VP1 plasmid (#22519) were purchased from Addgene (Cambridge, MA). VP1 expression plasmids encoding BKPyV genotypes Ia, II, III, IVc2 and were kindly provided by Dr Christopher Buck, National Cancer Institute (NCI), Bethesda, MD. The plasmid pEGFP-N1 (Clontech) was used as the reporter gene. Plasmids containing mutated VP1 sequences have been described previously were obtained by site-directed mutagenesis using the NEBase changer kit (New England Biolabs) (McIlroy et al., 2020).

### Labeled and non-labeled BKPyV VLP production

BKPyV virus-liked particles (VLPs) were prepared following the protocols developed by the Buck lab with slight modifications (Pastrana et al., 2012). Briefly, 1×10^7^ HEK 293TT cells were seeded in a 75 cm^2^ flask in DMEM 10% FBS without antibiotics, then transfected using Lipofectamine 2000 reagent (Invitrogen) according to manufacturer’s instructions. A total of 36 µg VP1 plasmid DNA was mixed with 1.5 mL of Opti-MEM I (ThermoFisher). 72 µL of Lipofectamine 2000 was diluted in 1.5 mL of Opti-MEM I and incubated for 5 min at room temperature prior to mixing with the diluted plasmid DNA. After 20 min at room temperature, 3 mL of DNA-Lipofectamine complexes were added to each flask containing pre-prepared 293TT cells.

Cells were harvested 48h post transfection by trypsinization and washed once in PBS then resuspended in one pellet volume (x µL) of PBS, then mixed with 0.4x µL of 25 U/mL type V Neuraminidase (Sigma). After 15 min at 37°C, 0.125x µL of 10% Triton X-100 (Sigma) was added to lyse cells for 15 min at 37°C. The pH of the lysate was adjusted by addition of 0.075x µL of 1M ammonium sulphate, or sodium bicarbonate if VLPs were to be fluorescence-labeled before ultracentrifugation, then 1 µL of 250 U/µL Pierce Nuclease (Pierce) was added to degrade free DNA. After 3h at 37°C, lysates were adjusted to 0.8M NaCl, incubated on ice for 10 min and centrifuged at 5000g for 5 min at 4°C. Supernatant was transferred to a new tube and pellet was resuspended in 2 pellet volumes of PBS 0.8M NaCl, then centrifuged. The second supernatant was combined with the first, then pooled supernatant was re-clarified by centrifuging. Cleared lysates containing BKPyV genotype I VLPs were labeled with either Alexa Fluor 555 or Alexa Fluor 647 and lysate containing MPyV VLP was labeled with Alexa Fluor 488 following Alexa Fluor Microscale Protein Labeling kit instructions (ThermoFisher).

Labeled or non-labeled lysate was layered onto an Optiprep 27%/33%/39% gradient (Sigma) prepared in DPBS/0.8M NaCl, then centrifuged at 175 000 *g* at 4°C overnight in an Sw55TI rotor (Beckman). Tubes were punctured with a 25G syringe needle, and ten fractions of each gradient were collected into 1.5 mL microcentrifuge tubes. 6.5 µL of each fraction was kept for SDS-PAGE to verify VP1 purity and determine peak fractions for pooling, then PBS 5% bovine serum albumin (BSA) was added to each fraction to get a final concentration of 0.1% BSA as a stabilizing agent. Peak VP1 fractions were pooled, then the VP1 concentration of each VLP stock was quantified by migrating 5µL of the stock on SDS-PAGE, then quantifying the VP1 band by densitometry using a standard curve constructed from a series of 4-fold dilutions of bovine serum albumin (BSA) starting at 5 µg/well. VLP morphology was confirmed at the electron microscopy facility at the Université François Rabelais, Tours.

### BKPyV Pseudovirus production

BKPyV Pseudovirus (PSV) particles were prepared following the protocols developed by the Buck lab with slight modifications (Pastrana et al., 2012). Briefly, cell preparation and transfection were performed similarly to BKPyV VLP production. However, instead of transfecting only VP1 plasmid, a total of 36 µg plasmid DNA consisting of 16 µg VP1 plasmid, 4 µg ph2b, 8 µg ph3b and 8 µg pEGFP-N1 was transfected into 293TT cells.

48h after transfection, producer cells were collected by trypsinization. The pellet was washed once in cold PBS then resuspended in 800 µL hypotonic lysis buffer containing of 25 mM Sodium Citrate pH 6.0, 1 mM CaCl_2_, 1 mM MgCl_2_ and 5mM KCl. Cells were subjected to sonication in a Bioruptor Plus device (Diagenode) for 10 minutes at 4°C with 5 cycles of 1 min ON / 1 min OFF. Type V neuraminidase (Sigma) was added to a final concentration of 1 U/mL and incubated for 30 min at 37°C. 100 µL of 1M HEPES buffer pH 7.4 (ThermoFisher) was added to neutralize the pH, then 1 µL of 250 U/µL Pierce Nuclease (Pierce) was added before incubation for 2 hours at 37°C. The lysate was clarified by centrifuging twice at 5000 *g* for 5 min at 4°C and PSV was purified in an Optiprep gradient as described for VLP production. After ultracentrifugation and fraction collection, 8 µL of each fraction was removed for qPCR and the peak fractions were pooled, aliquoted and stored at -80°C for use in neutralization assays.

For quantification of pEGFP-N1 plasmid, 8 µL of each fraction was mixed with 2 µL of proteinase K buffer containing 100 mM Tris-HCl pH 7.5 (ThermoFisher), 100 mM DTT (Sigma), 25 mM EDTA (Sigma), 1% SDS (Sigma) and 200 µg/ml proteinase K (Qiagen). This solution was incubated at 50°C for 60 min followed by 95°C for 10 min. Proteinase K extracts were diluted 80-fold in milliQ water and 1 µL was used for qPCR using Applied Biosystems 2x SYBR Green Mix (Applied Biosystems). Primers were CMV-F 5’-CGC AAA TGG GCG GTA GGC GTG-3’ and pEGFP-N1-R 5’-GTC CAG CTC GAC CAG GAT G-3’. Thermal cycling was initiated with a first denaturation step at 95°C for 10 min, followed by 35 cycles of 95°C for 15 sec and 55°C for 40 sec. Standard curves were constructed using serial dilutions from 107 to 102 copies of the pEGFP-N1 plasmid per tube.

### ELISA screening

Nunc MaxiSorp 96-well plates (Sigma) were coated with 50 ng of BKPyV VLPs in 50 µL PBS at 37°C overnight, then blocked with 5% powdered milk in PBS for 1h. Supernatants of transfected cells were used at 2 µL per well, and purified antibodies were assayed at 4-fold serial dilutions, starting at 10 µg/mL in 50 µL blocking buffer. Antibodies bound to VLPs were detected using goat anti-human IgG horseradish peroxidase-conjugated secondary antibody (Bethyl) diluted 1:5000 in blocking buffer. Washing was performed between each step with PBS 0.05% Tween-20 (Sigma). 50 µL TMB substrate (BD Biosciences) was added and the reaction was stopped by adding 50 µL 0.5M H_2_SO_4_. Absorbance was read at 450 nm in a TECAN Spark reader. The effective concentration 50% (EC50) was calculated using GraphPad Prism software.

### Neutralization assays

293TT and RS cells were seeded at a density of 1×10^4^ cells/well in flat bottom 96-well plates (BD Falcon) then allowed to attach at 37°C for at least one hour. BKPyV pseudovirus were diluted in the corresponding cell culture medium containing antibiotics, 0.1% BSA and 25 mM HEPES (ThermoFisher) to a concentration of 5×10^6^ EGFP-N1 copies/well for RS cells and 2×10^6^ EGFP-N1 copies/well for 293TT cells. Each produced antibody was added into PSV wells to make a series of 5-fold antibody dilutions starting at 500 ng/well (that is, 5 µg/mL). IgG-PSV mix was incubated at 4°C for 1h, then added onto plated 293T or RS cells. Plates were kept in a humidified 5% CO_2_ incubator at 37°C for 72h (293TT cells), or 96 hours (RS cells). Cells were washed once in PBS 0.5 mM CaCl_2_, 0.5 mM MgCl_2_ prior to fixing and staining in PBS 0.5 mM CaCl_2_, 0.5 mM MgCl_2_, 1% paraformaldehyde (EMS) and 10 μg/mL Hoechst 33342 (ThermoFisher). The number and percentage of GFP^+^ cells were quantified using a Cellomics ArrayScan VTI HCS Reader (ThermoFisher). Neutralization curves were constructed using GraphPad Prism software.

### Cloning, expression and purification of monoclonal antibodies

Monoclonal antibodies to BKPyV were produced as previously described (Devilder et al., 2018). The selected paired variable heavy (vH) and light chain (vL) sequences were sent to Eurofins for gene synthesis. vH and vL sequences were cut from the plasmid backbone by restriction enzymes prior to cloning into expression vectors containing constant regions of heavy chain (Cγ1 of IgG1) and light kappa chain (Cκ) or light lambda chain (Cλ). Cloned expression vectors were confirmed by Sanger sequencing, then plasmid Maxipreps were prepared.

Antibodies were first produced on a small scale to check specificity. The day before transfection, 1.5×10^4^ HEK 293A cells were seeded in 96-well plates in 200 μL DMEM supplemented with 1% Glutamax, 10% FBS. 125 ng of vH and 125 ng of vL expression vectors were diluted in 25 µL of 150 mM NaCl, then mixed with 0.5 µL transfection reagent jetPEI (Polypus) diluted in 25 µL of 150 mM NaCl. After 15 min incubation at room temperature, the complex was gently added onto pre-plated 293A cells. 16h post transfection, medium was replaced with serum-free medium Pro293a (Lonza) to avoid serum-Igs contamination. Cells were cultured for 5 days at 37°C in a humidified 5% CO_2_ incubator, then supernatants were harvested and centrifuged at 460 g for 5 min to eliminate cells and debris. The presence of antibody in supernatant was confirmed by ELISA using Affinity Purified goat anti-human IgG Fc Fragment (BD Biosciences) to coat plates and goat anti-human IgG horseradish peroxidase-conjugated (BD Biosciences) as secondary antibody.

Antibodies with confirmed VLP binding were scaled up for production and purification. Briefly, the day before transfection, 6×10^6^ HEK 293A cells seeded into 175 cm^2^ flasks were transfected with 10 µg of vH and 10 µg of vL expression vectors following jetPEI DNA transfection protocol. At 5 days post-transfection, supernatants were harvested and centrifuged at 460 *g* for 5 min to remove cells, then filtered using a 1.22 µm filter, then a 0.45 µm filter prior to purification.

Antibodies were purified using a 1 mL HiTrap rProtein A Fast Flow column (Sigma-Aldrich) on a fast protein liquid chromatography (FPLC) system (Bio-Rad). First, the protein A sepharose column was equilibrated with 20 mM pH 7.2 phosphate buffer. Filtered supernatant containing antibodies was loaded onto the column, then washed with 20 mM pH 7.2 phosphate buffer, and finally eluted with 0.1M pH 3 citrate buffer. 500 µL of each fraction was collected into tubes containing 1M pH 9 Tris buffer. Optical density was read at 280 nm on a spectrophotometer (Eppendorf) to determine peak fractions to pool. Pooled fractions were dialyzed in a Slide-A-Lyser cassette (ThermoFisher) with a 3.5K molecular weight cutoff against PBS overnight at 4°C with agitation, then sterilized by filtration at 0.2 µm. Antibody purity was verified by size-exclusion chromatography using Superdex200 Increase 10/300 GL column (GE Healthcare) following the manufacturer’s instructions.

### 10x Chromium Single-cell RNA sequencing with Immune profiling *B cell enrichment from PBMC*

B cells were isolated from frozen PBMCs using human B cell isolation kit II (Miltenyi Biotec) according to the manufacturer’s instructions. Briefly, frozen PBMC were thawed, counted and centrifuged then resuspended in 40 µL 1x PBS pH 7.2 buffer containing 0.5% BSA and 2 mM EDTA (Fluka), then labeled with a cocktail of biotinylated antibodies targeting non-B cells. Non-B cells were subsequently separated magnetically using the AutoMACS Pro Separator with “Deplete” program. Purified untouched B cells were collected and counted prior to labeling with antibodies and BKPyV VLPs.

### BKPyV-specific B cell staining and FACS sorting

Enriched B cells from 17 samples were resuspended in 100 µL of 1x PBS 1% BSA labeling buffer containing the following anti-human antibodies: anti-CD3-BV510 (diluted 1/20) (BD Pharmingen), anti-CD19-BV421 (diluted 1/50) (BD Pharmingen), BKPyV-gI-VLP-AlexaFluor555 (1.34 µg/mL), BKPyV-gI-VLP-AlexaFluor647 (0.54 µg/mL) and MPyV-VLP-AlexaFluor488 (1.34 µg/mL). Patient-specific and timepoint-specific TotalSeq-C oligonucleotide-labeled antibodies (BioLegend) were added into the mix as shown in Table 1, then incubated at 4°C for 30 min. Cells were washed three times in 13 mL of 1x PBS 1% BSA, centrifuged at 300 *g* for 5 min at 4°C. After the final wash, cells were resuspended in 1 mL of 1x PBS 0.04% BSA. 10 µL 7-AAD viability staining solution (BD Pharmingen) was added and incubated for 5 min in the dark. The 17 samples were pooled prior to sorting through a BD Aria FACS sorter (Becton-Dickinson). 1×10^5^ CD19^+^ B cells were sorted first, followed by BKPyV-specific B cells. The sorted cells were immediately used in the single-cell RNA seq procedure.

### Single-cell 5’ mRNA and VDJ sequencing

CD19^+^ B cells and BKPyV-specific B cells were loaded onto separate wells of a 10x Chromium A Chip. Subsequent steps were performed using 10x Chromium Single Cell 5’ Library & Gel Bead kit; 10x Chromium Single Cell 5’ Library Construction kit; 10x Chromium Single Cell 5’ Feature Barcode Library kit; 10x Chromium Single Cell V(D)J Enrichment kit, Human B Cell; 10x Chromium Single Cell A Chip kit and 10x Chromium i7 Multiplex kit according to the manufacturer’s protocols. Three libraries were prepared for each cell sample: a V(D)J enriched library, a 5’ gene expression library and a 5’ cell surface protein library. All libraries were quantified and verified using Quantus (Promega) and Caliper LabChip GX (LifeSciences). Libraries from the same cell sample were pooled at the ratio of 1 V(D)J Enriched : 5 5’ Gene Expression : 1 Cell Surface Protein and sequenced using an Illumina NextSeq500 system.

### B cell single-cell RNA seq data analysis

TotB and SpecB raw sequencing reads in FASTQ files obtained from Illumina sequencing were aligned with the human reference transcriptome (GRCh38) by the 10x Genomics cellranger count pipeline. Output matrices of filtered features of each B population were loaded separately into R version 3.5.1 using the Read10X function. Barcode matrices containing antibody-based hashtag oligo (HTO) count tables were added into each dataset, followed by a combination of parameters from the TotB and SpecB dataset to create a merged object. HTO counts were normalized by the centered-log ratio (CLR) method, followed by demultiplexing using the Seurat function HTODemux to trace back the cell’s origin. In a regular setting, doublets are removed; however, our experiments were designed to obtain double HTO-labeled cells, or doublets. Therefore, singlets and cells with wrong HTO combinations were eliminated from the dataset. Another quality control step was applied to filter out low quality cells which had the number of genes less than 500 and greater than 3000, the number of RNA counts less than 1000 and more than 15000. Cells with the percentage of mitochondrial genes more than 10% were also excluded. The remaining cells were then joined with the single-cell VDJ data to obtain merged B cells with productive pairs of heavy and light chain sequences. The merged B cells were used for further gene expression analysis.

Next, immunoglobulin variable genes were deleted from the dataset. To remove batch effects, the combined dataset was processed into the standard workflow of the Seurat Integration and Label Transfer pipeline (Stuart et al., 2019). The raw RNA counts were normalized in a log scale with a factor of 10.000 by default including the percentages of mitochondrial expression and the number of RNA counts as regression factors. Highly variable genes were identified by running FindVariableFeatures functions with the vst selection method, and the results were used as the input for principal component analysis (PCA). Performing RunPCA function returned 30 principal components (PCs) that were used to generate two-dimensional representations via RunUMAP function. Using the top 15 PCs together with the resolution of 0.1, cells were clustered by computing FindClusters and FindNeighbors functions. Clusters were visualized on a non-linear dimensional reduction plot UMAP.

### B cell single-cell VDJ data analysis

Sequencing FASTQ files were submitted to 10x Genomics cellranger vdj pipeline in the presence of the human VDJ reference (GRCh38) to perform sequence assembly and paired clonotype calling. For further analysis, data was extracted from the all_contig_annotations.json and filtered_contig_annotations.xls output files. Data were rearranged in R and OpenOffice Calc for repertoire analysis using the BRepertoire webtools (Margreitter et al., 2018), IMGT-V-Quest (Brochet et al., 2008) and Immunarch (www.immunarch.com). Firstly, clonotypes were identified in BRepertoire based on heavy chain V and J segment usage, then CDR3 homology initially using a Levenshtein distance of 20. Somatic hypermutations (SHM) were identified in heavy chain V-regions in IMGT-V-Quest, using the cell barcode as the sequence ID, then the number of non-synonymous heavy chain mutations was integrated to the dataframe, so that the proportion of IgM clones and mean number of SHM could be analyzed by clonotype using the dplyr package in R. Heavy and light chain V-gene usage, repertoire diversity and repertoire overlap were calculated and visualized using Immunarch, after substituting the clonotype ID numbers generated by BRepertoire for the CDR sequence used by Immunarch for clonotype identification. The Chao1 estimator for repertoire diversity was independently calculated using the vegan package in R, in order to determine the repertoire size using the different time points from individuals as independent observations.

## 4. Results

### Production and validation of labeled BKPyV VLPs

To generate VLPs, expression vectors containing VP1 sequences of either BKPyV genotype Ia or murine polyomavirus (MPyV) were transiently transfected into HEK 293TT cells. Cell lysates containing BKPyV were stained with either Alexa Fluor (AF) 555 or AF647, while MPyV lysate was stained with AF488. Labeled VLPs were then purified by ultracentrifugation through an Optiprep gradient and Optiprep was subsequently eliminated by repeated washes with 1x PBS in Amicon filters with a molecular weight cut-off of 100 kDa. Fluorescent VLPs had a homogeneous polyomavirus-like morphology in negative stain electron microscopy (Figure 1A) and SDS-PAGE showed that VLP stocks consisted of a single protein band corresponding to VP1, migrating at approximately 40 kDa for BKPyV and 42 kDa for MPyV (Figure 1B). Finally, no serological cross-reactivity was found between BKPyV VLPs and MPyV VLPs (Figure 1C), confirming that the binding of KTx patient antibodies to BKPyV VLPs to antibodies was specific, and that labeled MPyV VLPs could be used as a marker to eliminate non-BKPyV-specific B cells.

**Figure 1.**
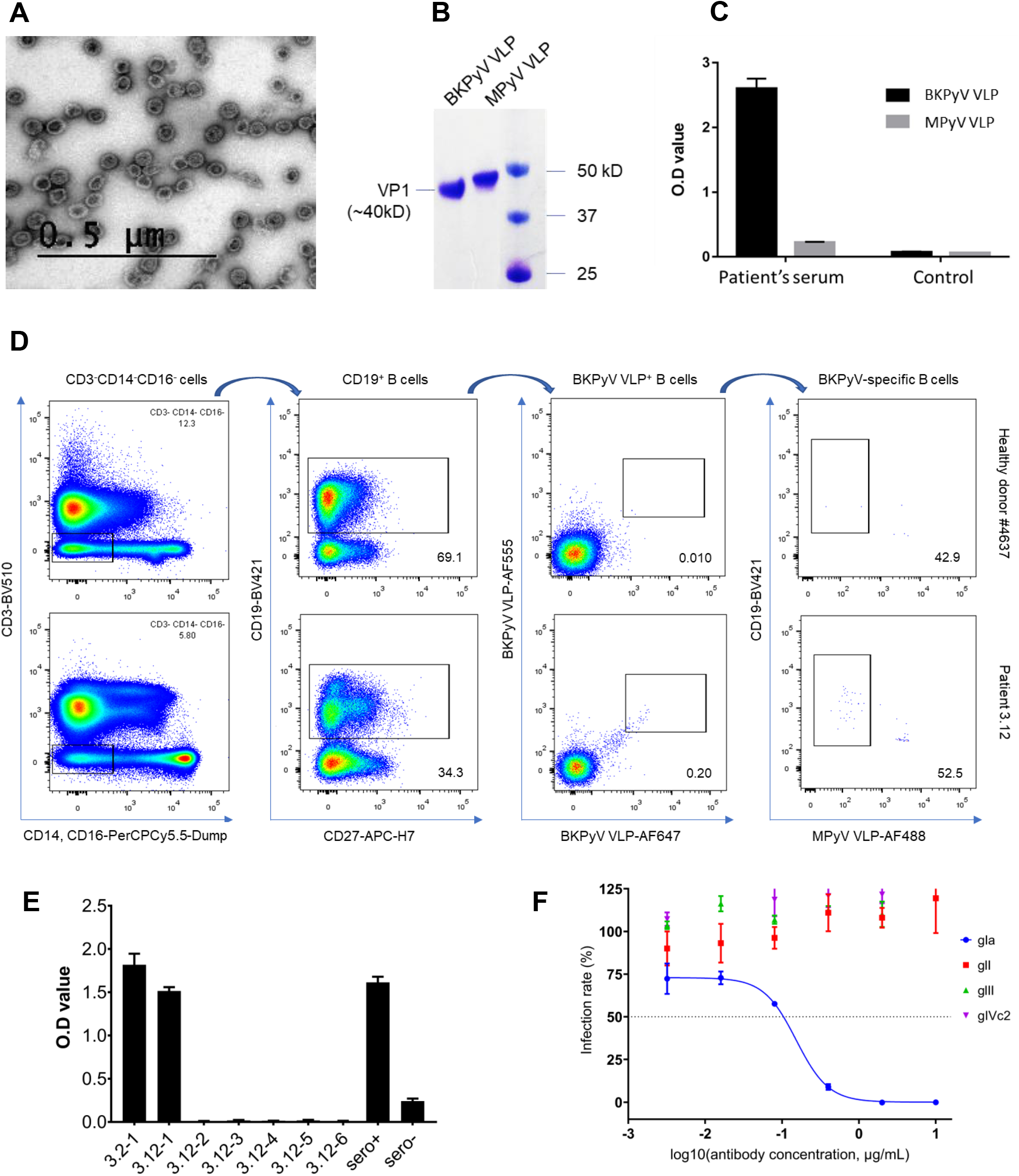
Fluorescence-labeled VLP characterization and isolation of BKPyV-specific B cells. (A) Negative stain electron microscopy of fluorescent VLPs. Bar = 0.5 µm. (B) Denatured labeled BKPyV VLP and MPyV VLP on SDS-PAGE protein gel. (C) IgG ELISA binding of serum from KTx recipient 3.1 and a seronegative KTx recipient to BKPyV and MPyV VLPs. (D) Binding of fluorescence-labeled BKPyV VLPs to B cells from healthy donors (upper panels) and seropositive KTx recipient with BKPyV reactivation (lower panels). From total PBMC, CD3^+^CD14^+^CD16^+^ cells were gated out, then CD19^+^ B cells were identified. A gate of AF555 and AF647 double-positive B cells were set and finally MPyV VLP^+^ cells were excluded. (E) Genotype I specific BKPyV binding and (F) neutralization of antibody 3.2-1.

Next we tested the ability of labeled BKPyV VLPs to target virus-specific memory B cells in different groups, including healthy donors, seronegative and seropositive KTx recipients post-transplant. 10-50 million frozen PBMC were thawed then stained with a cocktail of antibodies against CD3 (BV510), CD14, CD16 (PerCPCy5.5-Dump), CD19 (BV421), CD27 (APC-H7), BKPyV-gI VLP AF555, BKPyV-gI VLP AF647 and MPyV VLP AF488. FACS gating is shown in Figure 1D. Non-B cells (CD3^+^CD14^+^CD16^+^) were first removed, then CD19^+^ B cells were selected, and an analysis gate defined for AF555 and AF647 double-positive B cells. BKPyV-specific B cells were finally identified by a gate that excluded cells binding MPyV VLP. We observed that in 2 healthy donors, the percentage of BKPyV-specific B cells was extremely low, comprising no more than 0.01% of the CD19^+^ population (Figure 1D, one healthy donor shown: #4637). Similar proportions of BKPyV-specific B cells were detected in 2 BKPyV-seronegative KTx with no evidence of viral reactivation and in 1 seropositive patient with weak BKPyV reactivation (Supplementary Figure 1). In contrast, in 5 PBMC samples from 2 KTx recipients who were seropositive and experienced high BKPyV viruria and viremia, the percentage of BKPyV-specific B cells detected was always greater than 0.1%, at least 10-fold higher compared to the other groups (Figure 1D, patient 3.12). For the first attempt at isolating BKPyV-specific antibodies from patients 3.2 and 3.12, we sorted 14 and 49 single-B cells from which heavy and light-chain antibody sequences were amplified. We succeeded in reconstituting only 1 antibody from patient 3.2 (7%) and 6 from patient 3.12 (12%). An ELISA assay was conducted to identify antibodies that bind to BKPyV VLPs. Plates were coated with BKPyV-gIa VLPs, and incubated with generated antibodies. Among 7 antibodies tested, only 2 bound specifically to BKPyV VLP gIa (Figure 1E). To test neutralizing capacity, the 2 binders were incubated with BKPyV PSVs of each genotype, then inoculated onto 293TT cells. Only 1 binder specifically neutralized BKPyV PSV gIa (antibody 3.2-1, Figure 1F). These data showed that fluorescent BKPyV VLPs could be used as probes to sort circulating BKPyV-specific B cells.

### Sorting of BKPyV-specific B-cells from multiple KTx recipients

In order to obtain a sufficient number of BKPyV-specific B cells, we developed a strategy to process multiple frozen PBMC samples the same day, allowing us to obtain paired heavy and light chain antibody sequences that could then be screened for BKPyV-specific binding and neutralization (Figure 2A). Six KTx patients who had significant increases in neutralizing antibody titres following BKPyV reactivation (Supplementary Figure 2) were selected for this experiment. A total of approximately 270 million frozen PBMCs from 17 samples (at least two independent PBMC samples per patient) were thawed, then B cells were enriched from each sample by magnetic sorting. With a slight modification compared to the first staining strategy, enriched B cells were stained in parallel with antibodies against CD3-BV510, CD19-BV421, AF555 and AF647 labeled BKPyV VLP gIa, and MPyV VLP-AF488, in addition to two oligonucleotide hashtagged antibodies for each sample. These antibody-based oligonucleotide sequences enabled us to associate each cell’s RNAseq data to a specific PBMC sample during the demultiplexing analysis step (Supplementary Table 1). After separately processing and staining each PBMC sample, labeled B cells were pooled and FACS sorted. To study the total B cell repertoire, roughly 10^5^ CD19^+^ B cells were isolated first (TotB population), then the BKPyV-specific B cells were sorted from the remainder of the sample, yielding approximately 10^4^ sorted cells (SpecB population) (Figure 2B). Subsequent to FACS sorting, around 6×10^4^ TotB and 10^4^ SpecB cells were processed on the Chromium 10x platform in order to generate single-cell cDNA libraries for 1) heavy and light chain BCR sequences 2) 5’ gene expression profiles and 3) antibody hashtags identifying the sample. Libraries were pooled for the TotB and the SpecB sample, then sent for Illumina sequencing.

**Figure 2.**
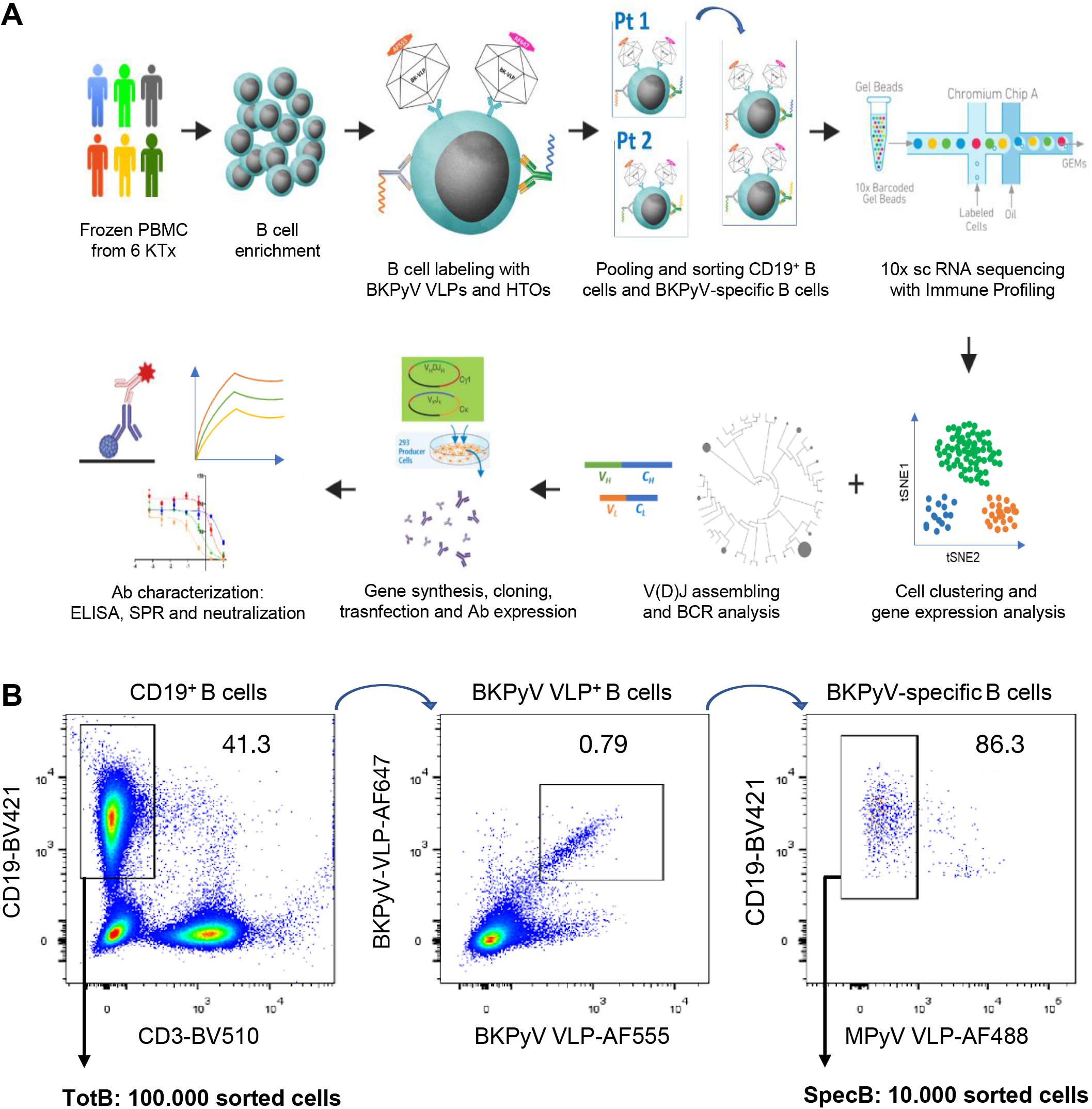
Experimental strategy for the analysis of BKPyV-specific B cell repertoire from multiple donors. (A) Strategy of the scRNA seq experiment and (B) FACS profile of pooled BKPyV-specific B cells.

### BCR repertoire of BKPyV-specific B-cells

Paired heavy and light chain BCR sequences were obtained for 2105 cells, with a very uneven distribution between patients. More than 1500 antibody sequences were derived from patient 3.1, while only 13 antibody sequences from patient 3.12 and only 12 from patient 3.3 were obtained. However, at least 50 BKPyV-specific BCR sequences were obtained from each of the other three patients (SpecB, Figure 3A), in addition to several hundred paired heavy and light-chain BCR sequences from circulating CD19^+^ B-cells from the same patients (TotB, Figure 3A). In all four patients with at least 50 BKPyV-specific antibody sequences, most BKPyV-specific B-cells carried Lambda light chains, whereas the total B-cell repertoire from the same patients was dominated by antibodies with Kappa light chains (Figure 3B). Only a minority of BKPyV-specific B-cells expressed IgA or IgG, even though the proportion of class-switched antibodies was higher than that observed in the corresponding total B-cell population for three out of the four patients (Figure 3C). In terms of V-gene usage, all four patients showed a greater than two-fold enrichment of IGVH4-39 in BKPyV-specific B-cells compared to total B-cells from the same patient, and in patients 3.1, 3.4 and 2.6, IGVH4-39 was the dominant heavy chain V-gene, present in at least 20% of BKPyV-specific B-cells (Supplementary Figure 3). In terms of light chain V-gene usage, no single V-gene stood out, although IGVL2-11 showed a greater than two-fold enrichment in three of the four patients.

**Figure 3.**
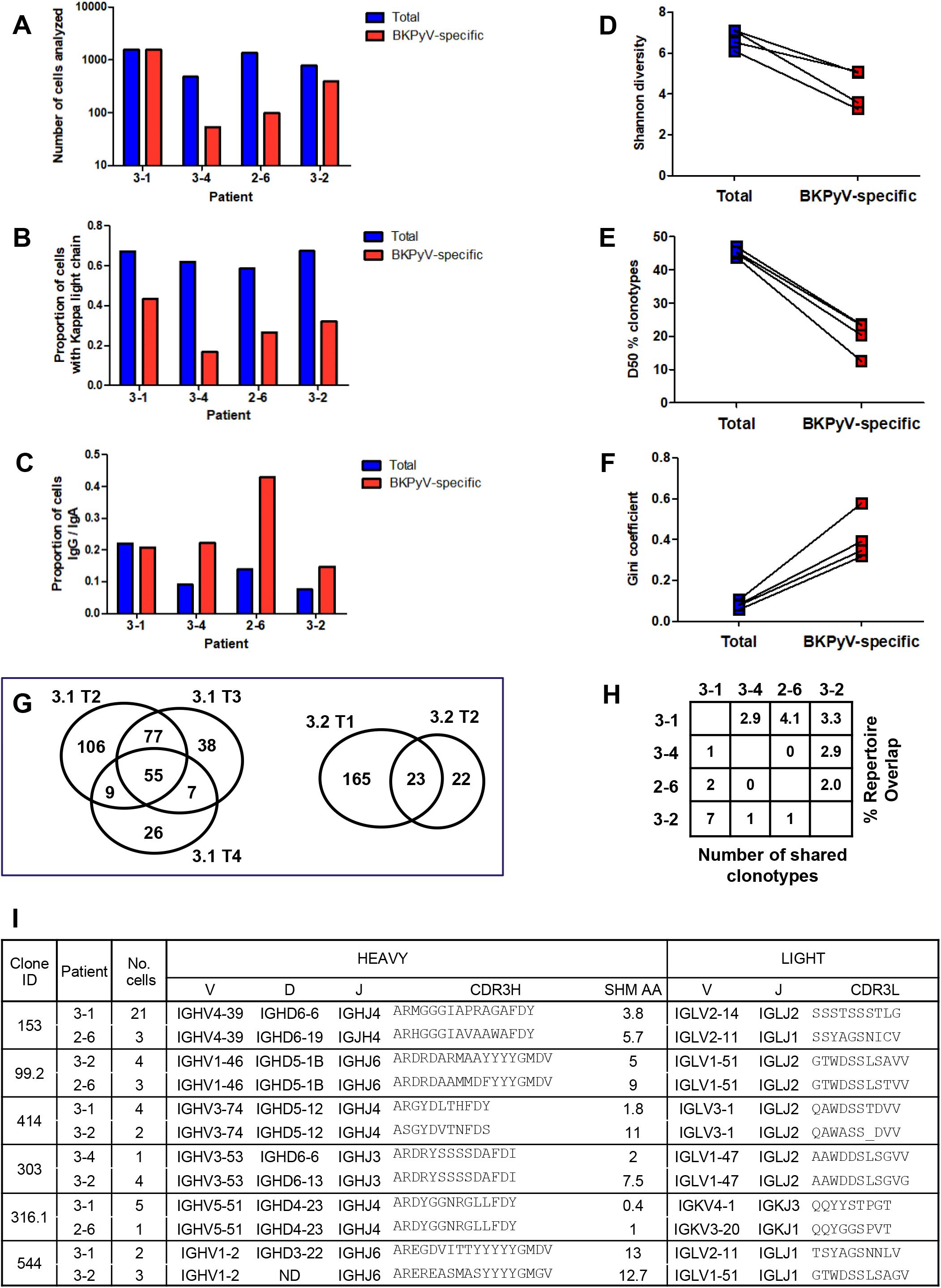
Features of BKPyV-specific BCR repertoire. (A) Number of cells with paired heavy and light chain antibody sequences obtained per patient from sorted BKPyV-specific (SpecB) and total (TotB) B cells. (B) Proportion of cells expressing Kappa light chain antibodies in SpecB and TotB datasets. (C) Proportion of cells expressing class-switched antibodies in SpecB and TotB datasets. (D-F) Shannon diversity, D50 index and Gini coefficient of clonotype distributions observed in SpecB and TotB datasets. Paired data represent cells from the same patient. (G) Venn diagram of B cell repertoire overlap in SpecB cells observed in independent PBMC samples from patients 3.1 and 3.2. Numbers represent the number clonotypes observed in each sector of the graph, independent of the number of cells per clonotype. (H) B cell repertoire overlap in SpecB cells between patients. The percentage overlap is calculated as the number of shared clonotypes divided by the number of clonotypes in the smaller dataset. (I) Sequence characteristics of selected “public” clonotypes.

Clonotype diversity, as measured by the Shannon diversity and the D50 index (Figure 3D-E) was significantly lower in BKPyV-specific B-cells compared to the total B-cell population, whereas the unevenness of the repertoire measured by the Gini coefficient (Figure 3F), was significantly higher. These characteristics are expected for B-cell repertoires biased toward recognition of a given antigen. Sufficient data were available for patients 3.1 and 3.2 to analyze the BKPyV-specific repertoire overlap in independent PBMC samples from the same individual. The BKPyV-specific repertoire overlap found in two samples from patient 3.2 was 51%, while for the three PBMC samples that provided sufficient data from patient 3.1, the average repertoire overlap was 68%. The availability of repeated samples from these two patients allowed us to calculate the α-diversity of the BKPyV-specific BCR repertoire using the Chao1 estimator. The total number of BKPyV-specific clonotypes was 437±25 in patient 3.1 and 599±99 in patient 3.2. Between patients, repertoire overlap was much lower – approximately 3% (Figure 3H) indicating that each patient’s BKPyV-specific BCR repertoire is mostly private, with only a few shared clonotypes between any two individuals. Since pooled cells were used to generate the BCR data in a single experiment, the possibility that errors in demultiplexing (for example, if two B-cells from the same clonotype in the same individual were erroneously assigned to two different patients) may have led to the spurious identification of shared clonotypes was addressed by inspecting the heavy and light chain V-(D)-J gene usage and CDR3 sequences in putative shared clonotypes (Figure 3I). In some cases, distinct light chain V-genes and/or heavy chain D-genes were used in the BCRs from different individuals, proving that these cells came from distinct clonal lineages. In other cases, despite identical heavy and light chain V-(D)-J gene usage, differences in CDR3 length or the number of heavy-chain non-synonymous SHM suggested a distinct clonal history for cells within the same clonotype assigned to different patients. We therefore concluded that these BKPyV-specific BCRs were indeed generated in distinct individuals, and represented genuine shared, or public clonotypes. Having validated the observed repertoire overlap between individuals, we estimated the size of the population level BKPyV-specific BCR repertoire as approximately 11400±3200 distinct clonotypes.

Next, we investigated the characteristics of IgM and IgG BKPyV-specific antibodies. As expected, the mean number of heavy-chain non-synonymous (NS) SHM was lower in B-cells expressing IgM or IgD compared to class-switched B-cells (Figure 4A). However, there were IgM outliers with very high numbers of SHM, and within clonotypes that contained both IgG and IgM antibodies, there was no difference in the mean number of SHM between IgG and IgM clones (Figure 4B). The relationship between class switching and SHM within clonotypes in BKPyV-specific B-cells was analyzed systematically in patients 3.1 and 3.2 by calculating the mean number of heavy chain SHM and the proportion of class-switched cells for each clonotype (Figure 4C). In both patients, a cluster of expanded IgM clones with low SHM that seemed to indicate a strong primary B-cell response were observed, as well as clonotypes with a progressively higher proportion of class switched B cells and a progressively higher number of SHM (Figure 4C), culminating in a cluster of clonotypes that were almost exclusively IgG, representing IgG memory B-cells (MBG). In addition, both patients also harbored a number of clonotypes with high levels of SHM that were predominantly IgM (upper-left quadrant, Figure 4C), which appeared to be IgM memory B-cells (MBM). These MBM included the dominant clonotype in patient 3.1, which had genotype I specific neutralizing activity when tested on 29TT cells and RS cells (Figure 5B).

**Figure 4.**
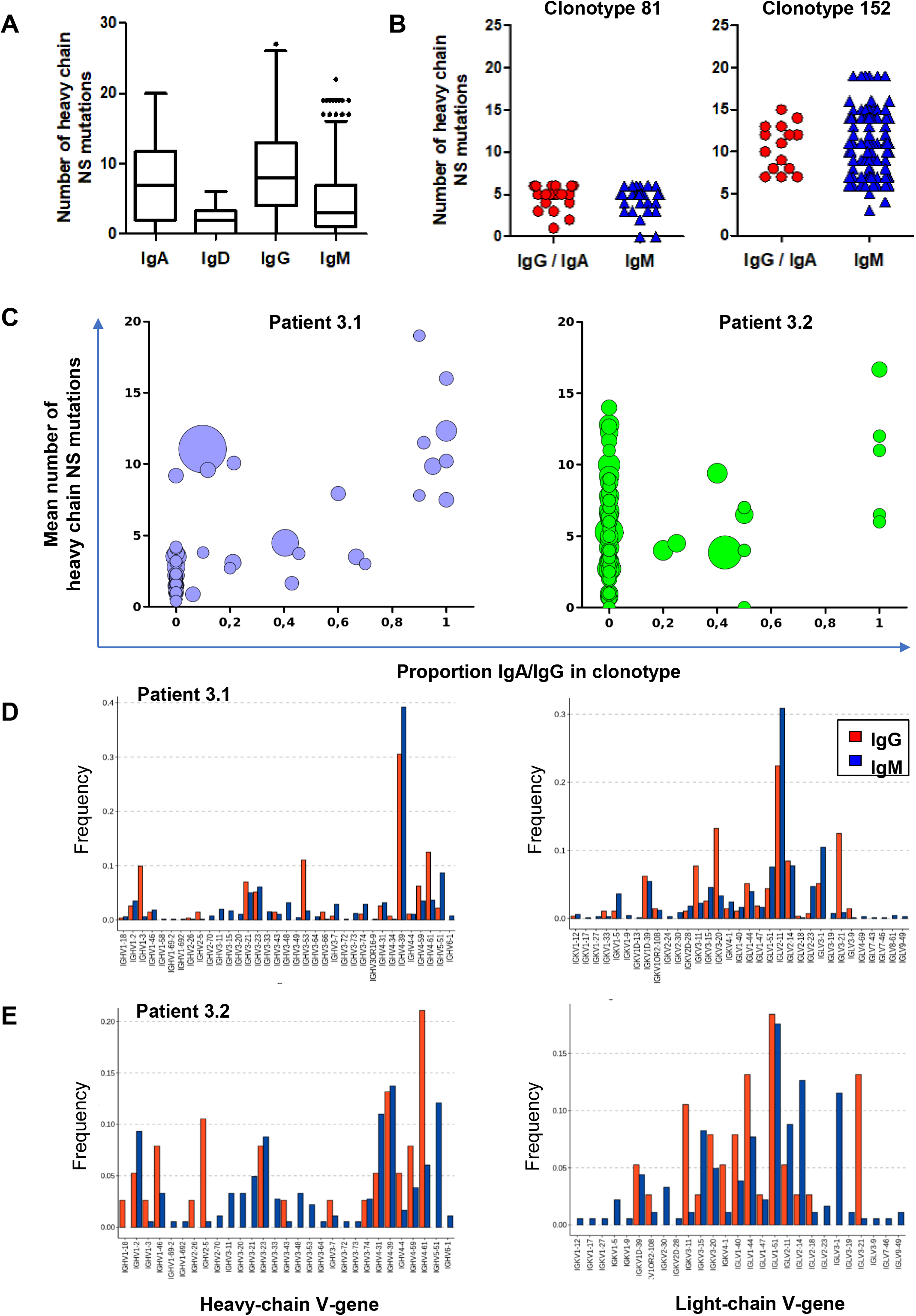
BKPyV-specific repertoire includes IgM memory B-cells with a BCR repertoire distinct from that found in IgG memory B-cells. (A) Number of heavy chain non-synonymous (NS) mutations in BKPyV-specific B cell repertoire as a function of antibody isotype. (B) Number of heavy chain NS mutations in class switched and IgM antibodies of the same clonotype. Two representative clonotypes are shown. (C) Mean number of heavy chain NS mutations per clonotype plotted as a function of the proportion of IgG/IgA cells within the clonotype. Bubble size is proportional to the number of cells within the clonotype, but the sizes are not normalized between patients. (D, E) Heavy and light chain V-gene usage in SpecB IgG and IgM antibodies with at least 5 heavy chain NS mutations in patient 3.1 (D) and patient 3.2 (E).

**Figure 5.**
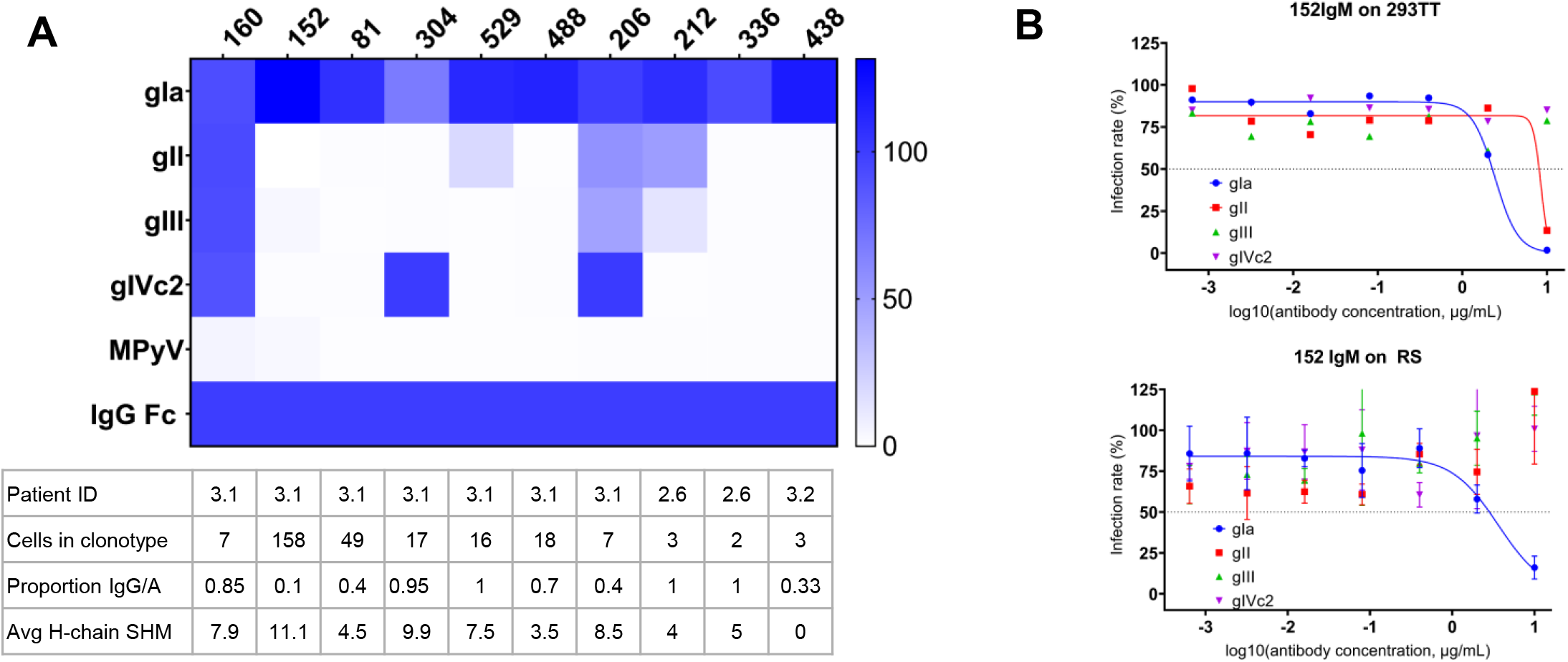
Specificity of selected antibodies in the SpecB dataset. (A) Binding properties of selected antibodies from the SpecB data set. Heatmap indicates ELISA OD450 values normalized to positive control binding to immobilized anti-IgG Fc antibody. (B) Neutralization properties of antibody 152 IgM in 293TT cells (upper panels) and RS cells (lower panels).

**Figure 6.**
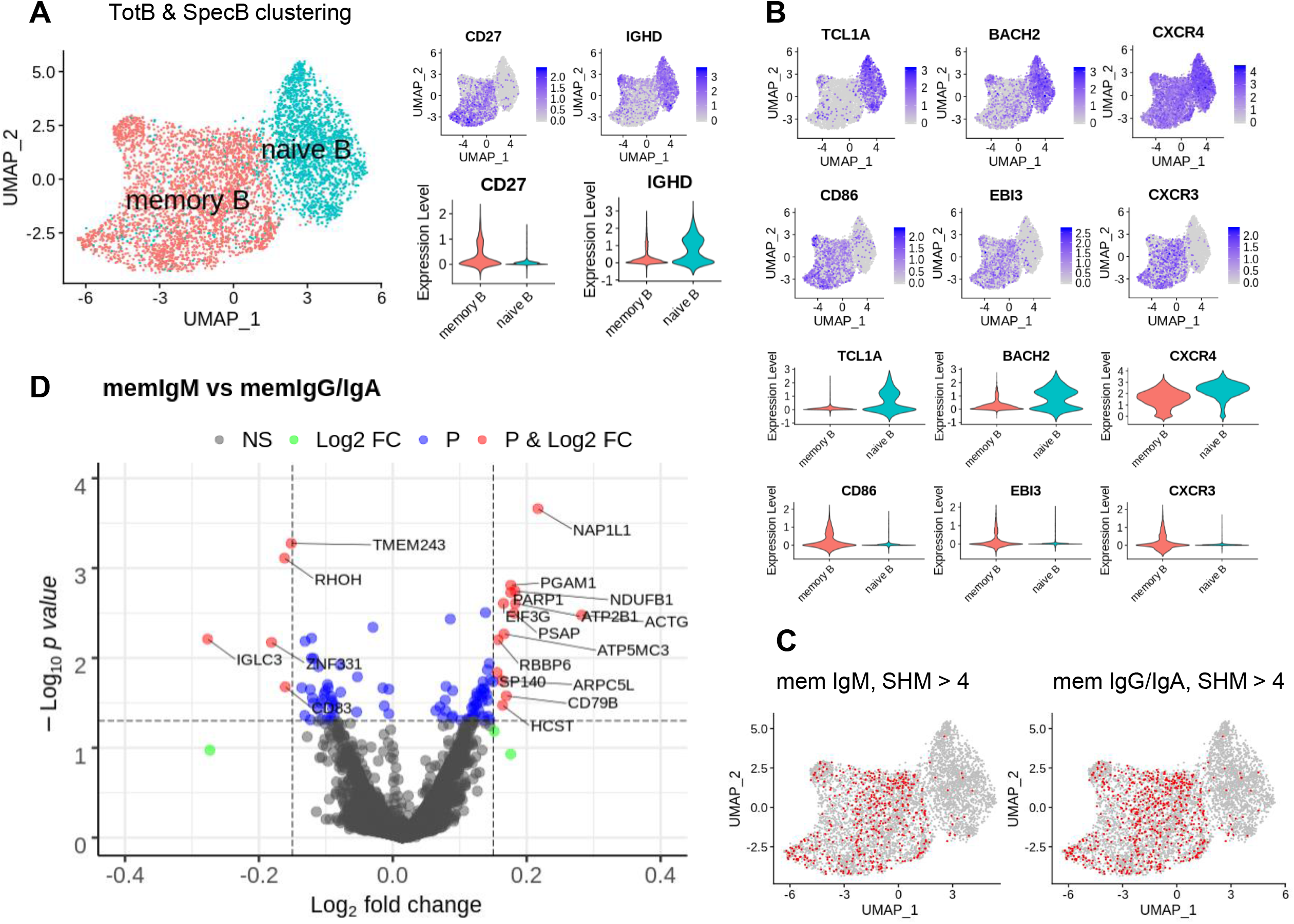
B cell clustering and differentially expressed gene analysis. (A) TotB and SpecB were merged, then clustered based on differential expression of genes (UMAP). The expression of CD27 marker and IgD are shown in feature plots (right, upper panels) and in violin plots (right, lower panels). (B) Expression levels of specific genes in naive and memory B clusters are displayed in feature plots (6 upper panels) and violin plots (6 lower panels). (C) IgM and IgA/IgG memory B cells with more than 4 SHM are projected on the clustering UMAP. (D) Volcano plot shows upregulated genes (right, red dots) and downregulated genes (left, red dots) in activated IgM memory B cells in comparison with activated IgA/IgG memory B cells. The *p*-value cut-off is 0.05.

We then compared V-gene usage in BKPyV-specific MBM and MBG, that is, cells expressing IgM or IgG antibodies with 5 or more NS heavy chain SHM. In both patients 3.1 and 3.2, BKPyV-specific MBM and MBG showed distinct patterns of heavy and light-chain V-gene use (Figure 4D and 4E). In patient 3.1, IGHV1-3, IGHV3-53, and IGHV4-61, and the light chain genes IGKV3-11, IGKV3-20 and IGLV3-21 were more frequent in MBG compared to MBM. A similar pattern was observed in patient 3.2, with IGHV2-5 and IGHV4-61, and IGKV3-11 and IGLV3-21 found predominantly in MBG. Conversely, IGHV5-51 and IGLV3-1 were preferentially used by BKPyV-specific MBM in both patients 3.1 and 3.2. The heavy-chain IGHV4-39 gene that showed the greatest enrichment in the overall BKPyV-specific BCR repertoire was found in both MBG and MBM. These results indicate affinity matured IgG and IgM antibodies specific for BKPyV use distinct BCR repertoires.

### Single-cell RNA seq profiling of BKPyV-specific B-cells

Single-cell RNA datasets of TotB and SpecB were integrated with V(D)J datasets to select only B lymphocytes with paired heavy and light chain sequences. After filtering out low quality and unwanted cells, we obtained and performed single-cell transcriptome profiling on 5450 cells including 3345 from TotB and 2105 from SpecB datasets. Using differential expression-based clustering and UMAP visualization, two distinct clusters were identified. We manually annotated these two clusters based on the expression of *CD27*, a reliable marker of human memory B cells, and IgD isotype (Figure 5A, UMAP). 3297 memory B cells were defined as IgD^-^CD27^+^; whereas, 2153 naive B cells were IgD^+^CD27^-^. Furthermore, the cluster of naive B cells showed high expression of *TCL1A, BACH2* as well as *CXCR4* (Figure 5B). *TCL1A* was demonstrated to be mainly expressed in naive B cells, and its expression diminishes proportionally to the maturation stages of B-cell development (Chong & Sciammas, 2011; Horns et al., 2020). Similarly, naive B cells at the early stage in the GC strongly express the transcription factor *BACH2* compared to memory B cells or plasma cells (Palm & Henry, 2019). The chemokine receptor *CXCR4* is upregulated in naive B cells, but downregulated in memory B cells and plays important roles in B-cell localization within the GC (Allen et al., 2004) and in controlling homeostasis of B cell compartments (Nie et al., 2004). Concerning the transcriptional signatures of memory B cells, elevated expression of genes associated with the activation of memory B cells and humoral responses was observed, such as the activation marker *CD86*, and the *EBI3* gene together with the chemokine receptor *CXCR3* (Figure 5B).

Next, we projected BKPyV-specific IgM and IgA/IgG memory B cells, which were defined as having greater than 4 SHM, on UMAP plots (Figure 5C). These cells were mainly found inside the cluster of memory B cells, and did not map to distinct sub-clusters within the memory B-cell population. As the BKPyV-specific antibody response was biased towards IgM^+^ B cells, we hypothesized that there might be specific genes that could distinguish IgM memory B-cells from class-switched memory B-cells. Analysis of differentially expressed genes between BKPyV-specific memory IgM and memory IgA/IgG using the negative binomial distribution test DESeq2 (Love et al., 2014) with a log2 fold change threshold of 0.15 revealed downregulation of 5 genes and upregulation of 14 genes (Figure 5D) in IgM memory B-cells compared to the expression of these genes in IgA/IgG memory B-cells. Among downregulated genes were *CD83*, which is a marker for mature dendritic cells and activated B cells during the GC reaction (Breloer & Fleischer, 2008; Krzyzak et al., 2016) and *IGLC3*, the lambda light chain constant region 3 gene. Among genes that were upregulated, we found *CD79B* which is associated with *CD79A* and membrane-bound immunoglobulin to form a mature BCR complex. Another B cell-involved gene that has a higher expression level in memory IgM subset is *PARP-1*. This gene acts in many important events during B-cell maturation (Yélamos et al., 2020). Overall, although we found various genes that were significantly differentially expressed (*p*-value less than 0.05), the levels of differential expression were small, indicating that the expression profiles of BKPyV-specific switched and unswitched memory B cells were highly similar.

## 5. Discussion

In order to study the BCR repertoire of BKPyV-specific B-cells in KTx recipients, we first validated that VLP purified from VP1-transfected 293TT cells had uniform polyomavirus morphology, and that AF555 and AF647 labeled BKPyV VLP could effectively stain susceptible cells (Supplementary Figure 4). Next, we tested whether labeled VLP could be used to specifically sort BKPyV-specific B cells. The frequencies of BKPyV VLP stained B cells exceeded 0.1% only in PBMC of KTx recipients who had experienced active BKPyV replication, and could not be reliably detected in PBMC from healthy donors. In contrast, Lindner et al. reported an average frequency of BKPyV-specific B-cells of 1/500 in three healthy blood donors using an unbiased screen of single B-cell cultures (Lindner et al., 2019). The frequencies of BKPyV specific B-cells that we observed may therefore be an underestimate, possibly due to the stringent FACS gating parameters that we applied.

Initial experiments sorting individual B-cells confirmed that BKPyV specific antibodies could be isolated from B cells that were specifically labeled with BKPyV VLP, although the yield of heavy and light chain sequences obtained was low. In order to increase the throughput of the analysis, we applied sc-RNAseq using the 10x Genomics Chromium platform, which has previously been used to obtain antibodies against influenza, HIV-1 (Setliff et al., 2019) and SARS-CoV2 (Cao et al., 2020; Shiakolas et al., 2020). The main technical obstacle was the requirement for approximately 10^4^ purified cells to load the Chromium chip: since the frequency of BKPyV-specific B cells was expected to be in the range 0.1 - 1%, this would require the processing of approximately 10^8^ PBMC. Each cryopreserved sample from a KTx recipient contained only 1 to 3×10^7^ PBMC, so similar to Cao et al. (Cao et al., 2020) we pooled samples from several patients in order to obtain a sufficient number of purified BKPyV-specific cells. However, we also used hashtag oligonucleotide labeled antibodies to demultiplex sequence data in order to assign each antibody to its PBMC sample. From 10^4^ sorted B cells, we obtained 2105 paired heavy and light chain sequences that were associated with a single patient sample, most of which were derived from patient 3.1. However, since at least 50 BKPyV-specific antibody sequences were obtained from three additional patients, it is possible to draw some conclusions concerning the BKPyV-specific BCR repertoire in KTx recipients.

Firstly, BKPyV-specific repertoires in all four patients were dominated by lambda light chain antibodies, with a clear inversion of the ratio of kappa/lambda light chain ratio between TotB and SpecB repertoires. Secondly, although significantly less diverse than the total B cell repertoires observed in the same patients, BKPyV-specific repertoires were highly varied in all four patients, with many different VH and VL genes contributing to the repertoire. Estimates of the overall clonotype diversity obtained in patients 3.1 and 3.2 indicated that an individual’s BKPyV-specific BCR repertoire consists of 400-700 distinct clonotypes. Analyzing each individual’s BKPyV-specific BCR repertoire as an independent sample of the γ-diversity led to an estimate of 8000-15000 distinct clonotypes, implying that each individual’s BKPyV-specific BCR represents approximately 5-10% of the available population-level repertoire. These estimates of repertoire diversity are based on two patients who controlled BKPyV replication, and may not be representative of patients with prolonged PyVAN, in whom virus neutralization escape occurs. As shown in Supplementary Figure 2, patients 3.3 and 3.4 displayed persistent high level BKPyV replication, and we have previously documented neutralization escape in these two patients (McIlroy et al., 2020; Peretti et al., 2018). Unfortunately, the low number of BKPyV-specific antibody sequences obtained from these two patients did not allow us to compare their BCR repertoires with those observed in patients 3.1 and 3.2, and further work will be required to systematically compare virus-specific BCR repertoires between patients with different virological and clinical outcomes.

Another potential bias in the estimation of repertoire size is the clonotype definition used: as clonotype definition becomes more stringent, the number of distinct clonotypes becomes larger, and the clonotype overlap between individuals becomes smaller. An extreme example of this is found in the clonotype definition in the 10x Genomics Loupe VDJ browser, which only classes antibodies as the same clonotype if they share identical heavy and light chain V- and J- gene usage, and share identical heavy and light chain CDR3 sequences. By this definition, even antibodies differing by a single CDR3 somatic hypermutation are classed as distinct clonotypes. On the other hand, using a less stringent clonotype definition introduces the risk of clustering unrelated antibodies together, in which case repertoire diversity would be underestimated. Some previous studies have used the Hamming distance as a clustering metric to group closely related antibody sequences (Jiang et al., 2013; Shlemov et al., 2017) and this has been shown to accurately define clonotypes within an individual’s BCR repertoire (Gupta et al., 2017). However, in the present work, we defined clonotypes on the basis of heavy chain CDR3 nucleotide sequence using the Levenshtein distance, which can account for insertions and deletions, rather than the Hamming distance, which cannot. The reasoning behind this choice was to enable us to group antibodies with the same specificities into clonotypes, even if they were expressed by B cells with distinct clonal histories, in which case CDR3 length could differ. Furthermore, because only heavy chain sequences contributed to the clonotype definition, we were able to use light chain sequences as an independent verification of clonotype validity. Within individuals, antibodies defined as the same clonotype based on the heavy-chain sequences also shared the same light chain V- and J- gene usage and CDR3 sequence, and as shown in Figure 3I when shared clonotypes were found between individuals, light chain CDR3 sequences were generally also very similar. These observations suggest that our strategy for clonotype definition was valid, although we cannot rule out the possibility that some clonotypes (for example clonotype 544, Figure 3) may have placed two distinct antibodies within the same cluster. Another concern is whether all the antibody sequences in the SpecB dataset were in fact specific for BKPyV, particularly since only two of seven antibodies expressed from sorted B-cells were BKPyV-specific in preliminary experiments (Figure 1E). Significantly, six of the initially screened antibodies came from cells sorted from patient 3.12, from whom we only obtained 13 antibody sequences in the scRNAseq experiment. It therefore seems likely that BKPyV-specific B-cells were relatively infrequent in this patient, which could explain the low ratio of BKPyV-specific B-cells observed in our initial experiments. On the other hand, all 10 of the antibodies that we expressed from the SpecB dataset bound specifically to BKPyV VLP, even when the antibody clonotype was represented by only two or three cells (Figure 5). We therefore have confidence that clonotypes observed at least twice in the SpecB dataset are indeed BKPyV-specific.

The third reproducible feature of the BKPyV-specific BCR repertoire was the predominance of IgM antibodies, which confirms previous observations in healthy donors (Lindner et al., 2019). MBM have also been described in the context of humoral responses to Gram negative bacteria (Rollenske et al., 2018), *Plasmodium falciparum* (Tan et al., 2018; Yilmaz et al., 2014) and more recently, SARS-CoV2 (Newell et al., 2020). In mice, MBM persist longer than MBG (Pape et al., 2011). If the same were true in humans, this might explain the contribution of MBM to the BKPyV-specific BCR repertoire in KTx recipients. Primary BKPyV infection occurs during childhood, and although the virus persists in the kidney, serum antibodies wane over time, indicating that the humoral response is not persistently stimulated. The KTx recipients studied here were all more than 50 years old, so over the decades that separated primary BKPyV infection from the superinfection after KTx, MBM may have become progressively more frequent compared to MBG, leading to the dominance of MBM that we observed in the SpecB dataset. However, virus-specific MBM were observed to be short lived in Yellow-Fever vaccine recipients, and in two convalescent patients recovering from an acute flavivirus infection (Wec et al., 2020). It is therefore not clear whether the relative longevity of MBM compared to MBG observed in the mouse is recapitulated in humans.

Another important feature observed in BKPyV-specific MBM, is that they expressed a distinct BCR repertoire compared to that found in BKPyV-specific MBG. Repertoire differences between MBM and MBG are well documented in circulating B-cell subsets (Bagnara et al., 2015; Wu et al., 2010), but to the best of our knowledge, the present work is the first time that IgM and IgG repertoires have been compared in antibodies specific for the same antigen. Antibody responses dominated by IgM are characteristic of T-independent antigens (Allman et al., 2019), and indeed, the humoral response to mouse polyomavirus is T-independent (Guay et al., 2009; Szomolanyi-Tsuda & Welsh, 1996). Polyomavirus capsids are rigid structures presenting multiple copies of the same epitope in an ordered array, which is similar to the structural properties of the TI-2 antigens. However, this property alone cannot explain why distinct B-cell clones directed against the BKPyV capsid differentiate towards either MBG or MBM. One explanation for this observation could be that IgM and IgG antibodies recognize distinct epitopes on the capsid. In this regard it is tempting to speculate that 5-fold and 6-fold symmetry in the distribution of certain epitopes on polyomavirus capsids may favour IgM antibodies. If this is the case, then one would expect that IgM will make a significant contribution to the B-cell memory repertoire specific for other non-enveloped viruses with icosahedral capsids. Structural studies will be required to test this hypothesis. Alternatively, the distinct repertoires observed in BKPyV-specific MBM and MBG could reflect the different processes of B-cell activation and differentiation occurring in different clonal lineages, with B-cells that respond to BKPyV capsids in an extrafollicular activation process differentiating into MBM, while those clones that enter germinal centres give rise to MBG lineages. The lower expression of *CD83* that was seen in the MBM subset is consistent with this model, as is the observation that many BKPyV-specific IgM clonotypes had little SHM, even though the expression profile of these cells placed them in the memory B-cell population. As a consequence, the MBM repertoire would be more diverse and more extensive than that found in MBG, but perhaps less effective in the antiviral response in terms of specificity or neutralization potency.

In addition to repertoire differences, we also studied the gene expression profiles of BKPyV-specific MBM from MBG. Both types of cells were found predominantly within the memory B-cell cluster defined on the UMAP plot, and did not appear to define specific sub-clusters within the memory B-cell population (Figure 5A). Accordingly, differences in gene expression between BKPyV-specific MBM from MBG were minor, concerning only a handful of genes. Interestingly, the expression of *PARP-1* was found to be slightly increased in the MBM subset. Affinity maturation of antibodies is characterized as somatic hypermutation and class-switch recombination which require AID. However, AID activity is restricted in the presence of PARP-1 at Ig loci due to its mutagenic repair function, leading to a decrease of the number of somatic hypermutations (Paddock et al., 2010; Tepper et al., 2019). This can be an explanation for the lower mean number of non-synonymous SHM in the MBM subset compared to that in the MBG subset in our dataset, even though they were both activated memory B cells. Furthermore, *PARP-1*-deficient mice stimulated by T cell-independent antigens expressed predominantly IgG1 and IgG2b antibodies (Ambrose et al., 2009; Galindo-Campos et al., 2019; Robert et al., 2009). Similarly, inhibition of *PARP-1* enhanced the abundance of class-switched immunoglobulins (Shockett & Stavnezer, 1993). These findings suggest the involvement of *PARP-1* in suppressing class-switching in MBM. Further investigations are required to confirm this hypothesis.

## Data Availability

The single cell RNAseq dataset will be supplied by the authors on request.

## Acknowledgments

The authors would like to thank Chris Buck for advice in setting up VLP and PSV purification protocols, Jacques LePendu and Antoine Touzé for discussions at several stages of this work, and Antoine Touzé for help with electron microscopy.

