## SupplementaryTable_Figures for "The humoral response to BK polyomavirus in kidney transplant recipients is dominated by IgM antibodies that use a distinct repertoire compared to IgG against the same antigen"

Supplementary Table 1:

Combinatorial sample barcoding

| Patient | Patient-specific antibodies | Timepoint-specific antibodies | | | |
|---|---|---|---|---|---|
| | | **Timepoint 1** | **Timepoint 2** | **Timepoint 3** | **Timepoint 4** |
| 2.6 | Hashtag 1 | anti-CD11a | anti-CD18 | anti-CD45 | |
| 3.1 | Hashtag 2 | anti-CD11a | anti-CD18 | anti-CD45 | anti-CD20 |
| 3.12 | anti-HLA-ABC | anti-CD11a | anti-CD18 | | |
| 3.2 | anti-HLA-DR | anti-CD11a | anti-CD18 | | |
| 3.3 | Hashtag 4 | anti-CD11a | anti-CD18 | anti-CD45 | |
| 3.4 | Hashtag 5 | anti-CD11a | anti-CD18 | anti-CD45 | |

* *Barcoded antibodies against human surface antigens with TotalSeq C oligonucleotides, specifically compatible with the Single Cell 5' workflow of 10X Genomics.*

Supplementary Figure 1:

Binding of fluorescence-labeled BKPyV VLPs to B cells from healthy donor, seronegative KTx, seropositive KTx

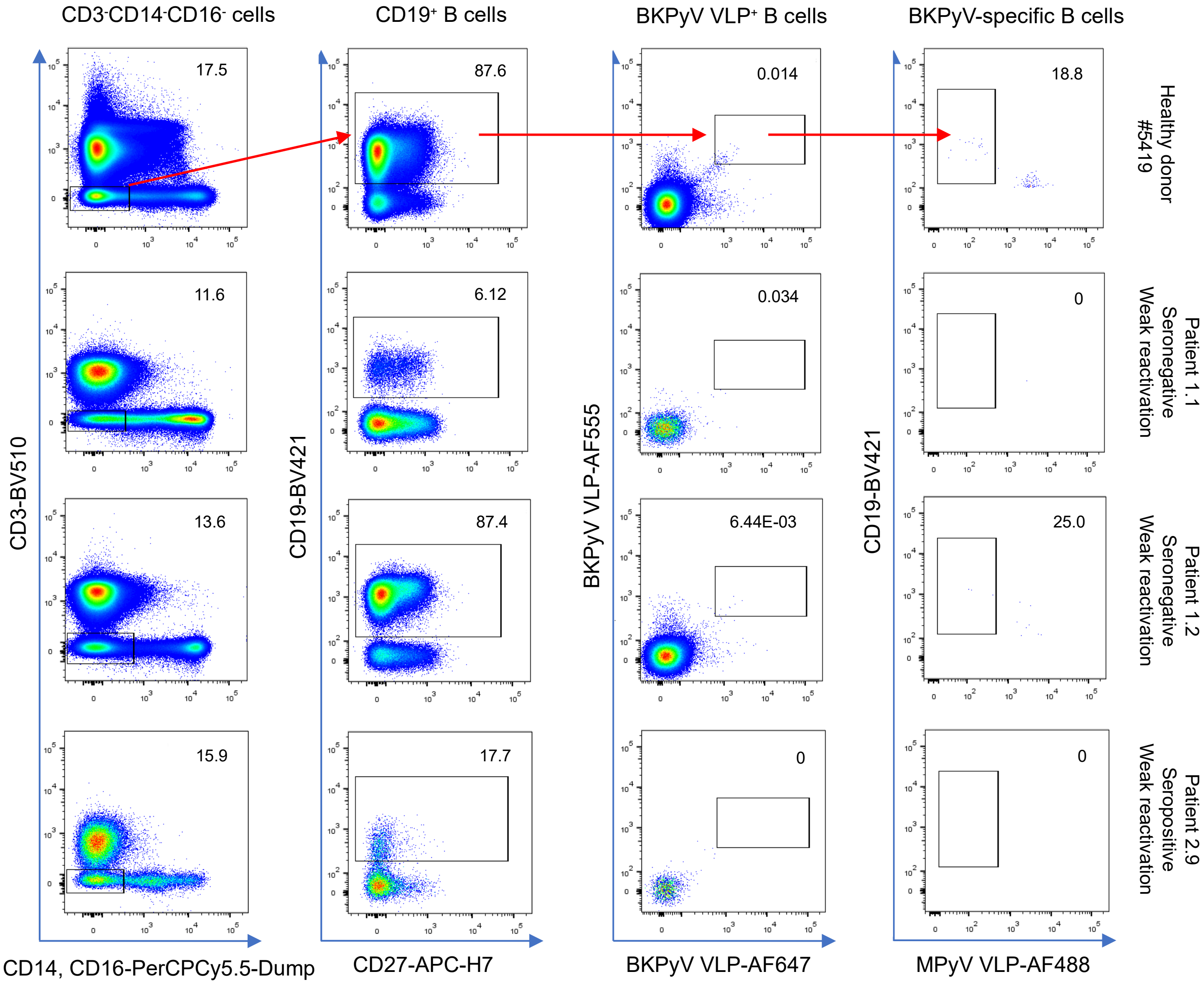

Supplementary Figure 2- Viral load, neutralizing antibody titres, and summary clinical data of patients selected for scRNA seq experiment

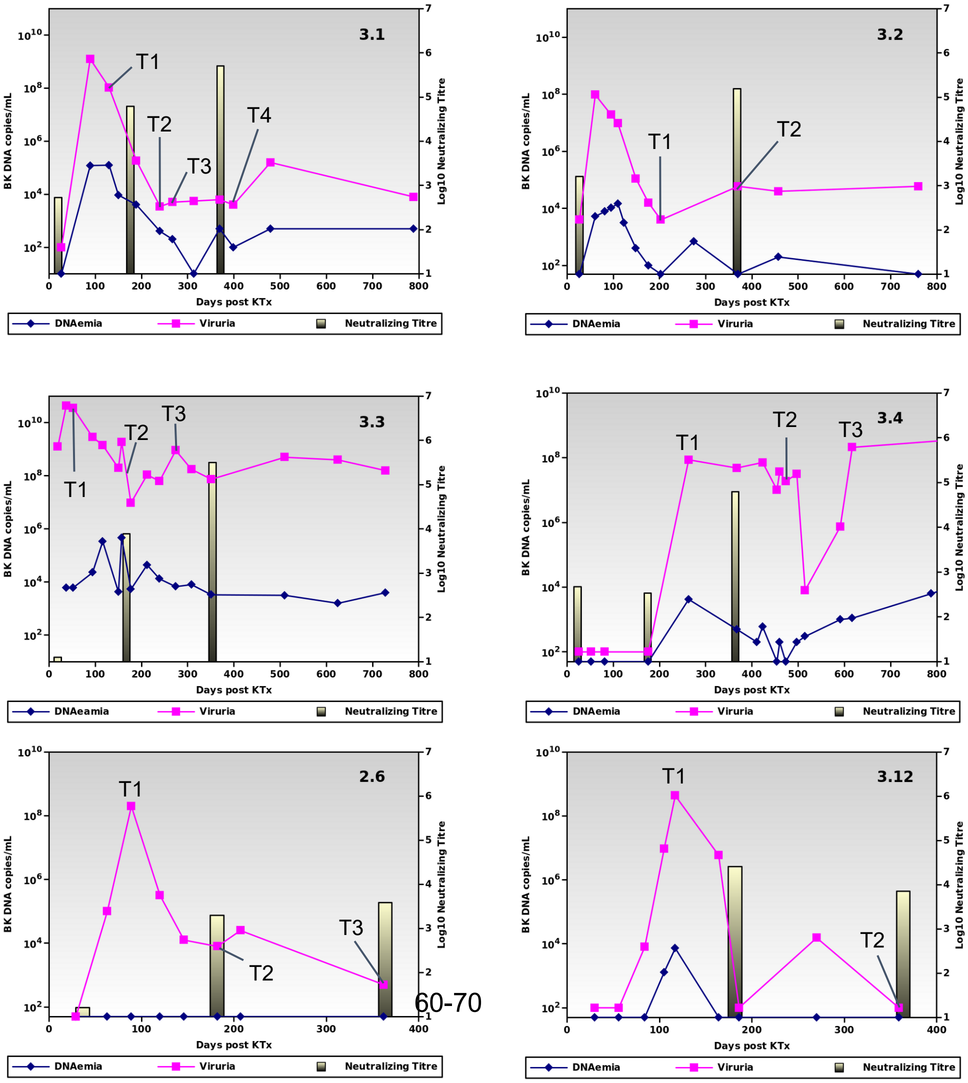


| Patient | Sex | Age at KTx | BKPyV genotype |
|---|---|---|---|
| 3.1 | F | 60-70 | Ib2 |
| 3.2 | M | 60-70 | Ib2 |
| 3.3 | M | 60-70 | IV |
| 3.4 | M | 60-70 | Ib2 |
| 2.6 | F | 50-60 | Ib1 |
| 3.12 | M | 50-60 | Ib2 |

### Supplementary Figure 3:

#### Heavy and light chain v-gene usage in total and BKPyV-specific B-cells

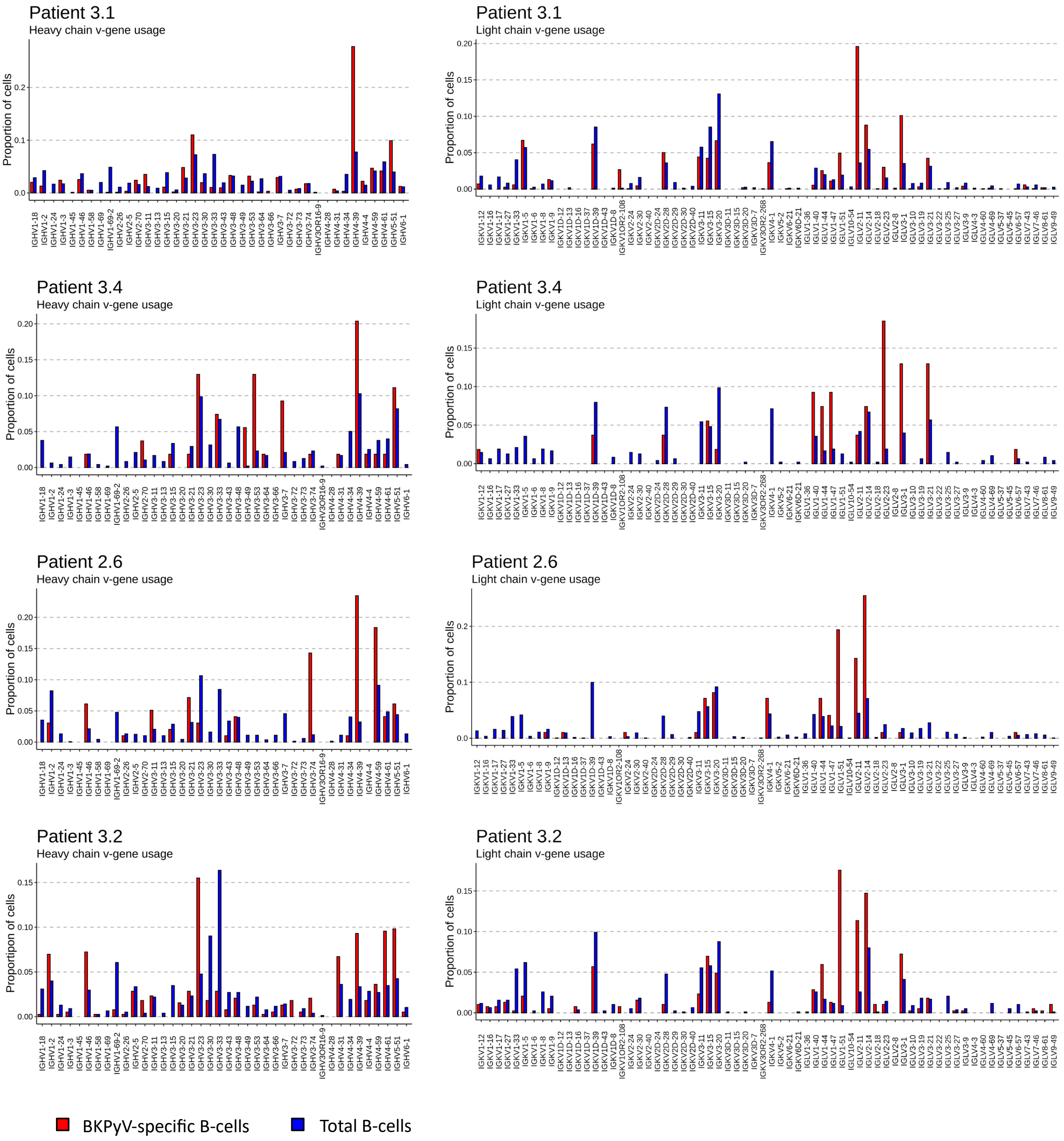

### Supplementary Figure 4:

#### Binding of fluorescence-labeled BKPyV VLPs to susceptible 293TT cells

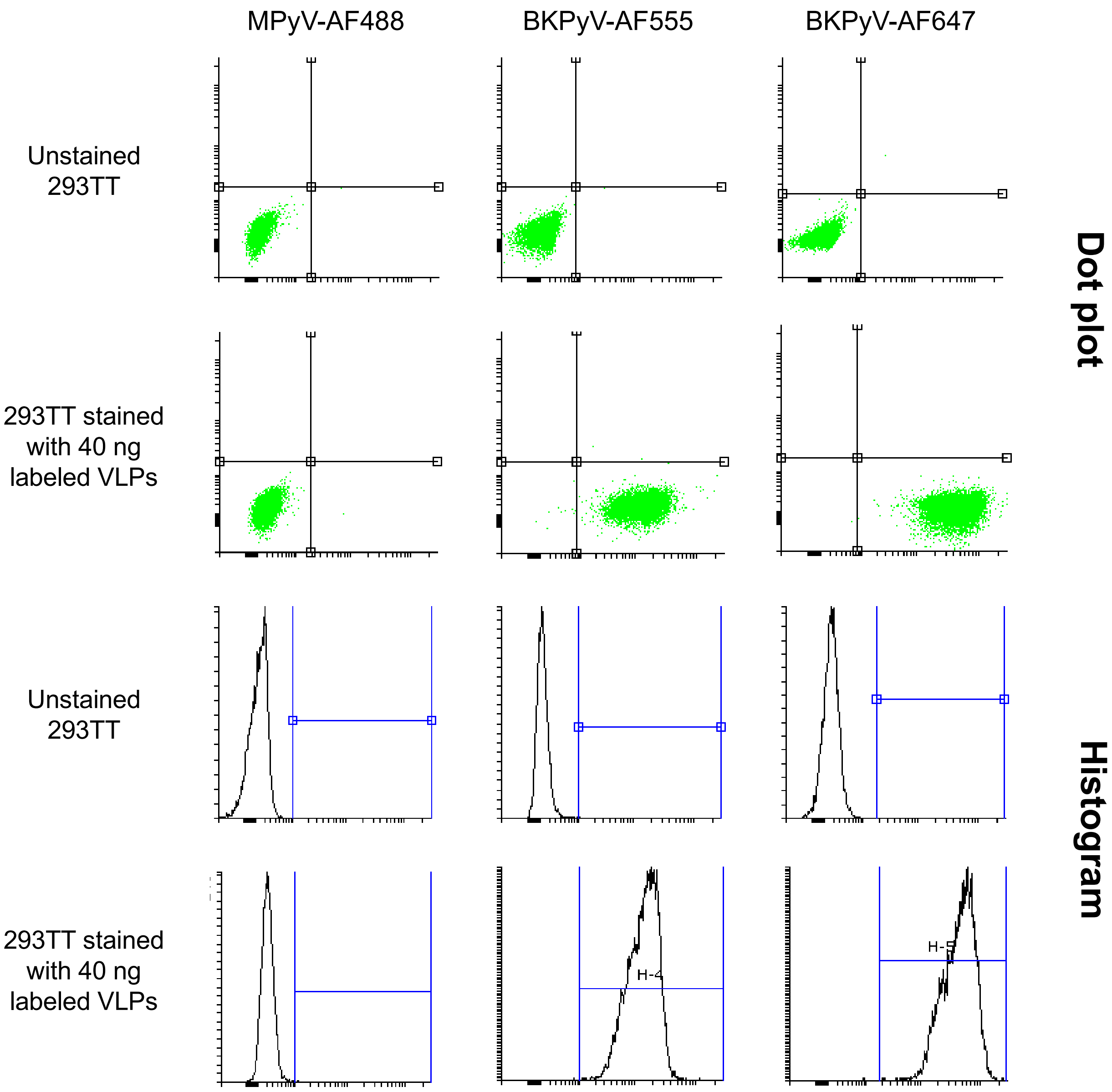
